# Methods used to Account for Concurrent Analgesic Use in Randomized Controlled Trials of Interventional Pain Treatments: A Meta-Epidemiologic Study

**DOI:** 10.1101/2024.11.02.24316637

**Authors:** Balaji V Sridhar, Andrew Humbert, Adam Babitts, Carina Staab, Clinton J. Daniels, Malka Dhillon, Patrick Heagerty, Joshua Goldenberg, Mark Jensen, Pradeep Suri

## Abstract

**Objective:** The modest effect sizes of most pain treatments make it essential that randomized controlled trials (RCTs) use methods that clearly define treatment effects of interest and consider the role of concurrent treatments. This study aims to determine how frequently concurrent analgesic use is reported in interventional pain RCTs and how accounting for analgesic use can affect pain intensity outcomes.

**Design:** Meta-epidemiologic study.

**Methods:** We conducted a study of concurrent analgesic use among RCTs from a recent systematic review of non-surgical interventional pain treatments (n= 37). We calculated the prevalence of different methods used to report concurrent analgesic use. We performed meta-analyses to compare treatment effects on pain intensity with vs. without accounting for concurrent analgesic use via a novel quantitative composite outcome, the “QPAC_1.5_.”

**Results:** About half of interventional pain RCTs reported concurrent analgesic use, but only one RCT directly accounted for concurrent analgesic use in their pain intensity outcome. Analyses accounting for concurrent analgesic use using the QPAC_1.5_ substantially increased the estimated treatment effect of interventions on pain intensity by an average of −0.45 numeric rating scale points (95% CI −0.76 to −0.14; p<0.001), as compared to analyses that did not adjust for analgesic use.

**Conclusion:** Concurrent analgesic use is sometimes reported in interventional pain RCTs, but rarely accounted for when examining treatment effects on pain intensity. Accounting for concurrent analgesic use changes the treatment effect of interest to remove differential analgesic rates and has the potential to significantly affect estimates of effect sizes on pain treatments.

## INTRODUCTION

Pain affects 100 million adults in the United States (US) and costs up to $635 billion annually.^1,2^ Many interventional pain treatments have been developed to mitigate the negative impacts of pain. Randomized controlled trials (RCTs) provide the strongest level of evidence to support pain treatment effectiveness.^3,4^ However, the modest effect sizes of most current pain treatments make it essential that RCTs use the most rigorous analytic methods to reduce the likelihood that meaningful treatment effects are obscured by other factors.

The use of analgesics following randomization may be an important factor to account for in interventional pain RCTs. Differential post-randomization analgesic use between intervention and control groups in an RCT may reflect participants’ efforts to self-manage pain intensity after receiving the randomized treatment, and such efforts may decrease the magnitude of treatment effects on pain intensity in an RCT if not accounted for. ^5,6^ Our observation is that concurrent analgesic use is not well-reported in many RCTs evaluating pain treatments and, even when reported, is not explicitly considered when defining or estimating treatment effects on pain intensity. Consistent with this, a systematic review of analgesic treatment RCTs for back pain and neuropathic pain found that about half of trials allowed study-specified “rescue” analgesic use and/or participants to continue to use analgesics other than the randomized treatment.^7^

A recent study by Suri et al. found that ignoring concurrent analgesic use in the analysis of pain RCTs may lead to substantial underestimation of treatment effects on pain intensity and may affect power.^5^ Simple methods of accounting for concurrent analgesic use, such as analyzing analgesic use as a binary outcome or adjusting for it as a covariate, decreased power.^5^ Suri et al. proposed a new measure to account for concurrent analgesic use: the Quantitative Pain and Analgesic Composite outcome (QPAC_1.5_). The QPAC_1.5_ is a composite measure that combines pain intensity and concurrent analgesic use into a single 0 to 10 numeric rating scale (NRS) of pain intensity. Prior work has suggested that concurrent analgesic use has an average impact on perceived pain intensity of 1.5 NRS points ^6^. Accordingly, the QPAC_1.5_ attributes a 1.5-NRS-point weight to concurrent analgesic use when approximating a research participant’s counterfactual pain NRS rating that *would have been* reported in the absence of analgesic use, by adding 1.5 NRS points to a participant’s conventional pain intensity NRS rating if the participant is taking analgesics at the time of pain intensity reporting. While the QPAC_1.5_ applies a 1.5-NRS-point weight to concurrent analgesic use, the approach is flexible, and could instead apply a different weight if appropriate for a given clinical population. The QPAC_1.5_ has the same range and interpretation as the conventional pain intensity NRS, facilitating easy comparisons between the two measures.^5^ Statistical simulations and analyses of trial data have found that the QPAC_1.5_ may increase power in pain RCTs relative to analyses that look only at the total effects of treatment on pain intensity without removing impacts on pain intensity that occur due to differential analgesic use. ^6^

To our knowledge, no prior studies have evaluated how often interventional pain RCTs report on concurrent analgesic use or examined whether the analysis methods used accounted for analgesic use when analyzing pain intensity outcomes. Similarly, no prior meta-analyses have compared treatment effects on pain intensity with and without the use of methods to account for post-randomization concurrent analgesic use. Therefore, we conducted a meta-epidemiologic study of concurrent analgesic use reporting among RCTs included in a 2021 comprehensive systematic review of emerging non-surgical interventional procedural treatments for pain. ^8^ The primary study aims were to examine (1) the prevalence of reporting concurrent analgesic use among included RCTs, and (2) the methods used to account for concurrent analgesic use. A secondary aim of the study was to investigate the impact of accounting for post-randomization concurrent analgesic use on estimated treatment effects on pain intensity, using the QPAC_1.5_.

## METHODS

### Data Source

Before any study-related activities began, the study protocol and analysis plan were registered on the Open Science Framework (https://osf.io/syug3/).^9^ We conducted a meta-epidemiologic study of interventional pain treatment parallel-group RCTs identified from a 2021 systematic review by Chou et al., to determine the prevalence of reporting concurrent analgesic use and how analgesic use was accounted for in analyses of pain intensity. This high-quality systematic review followed the methods suggested in the *Agency for Healthcare Research and Quality Methods Guide for Effectiveness and Comparative Effectiveness Reviews,* which was developed for the Evidence-based Practice Centers.^8^ The review methods were determined *a priori* using a protocol developed through a process that included public input and was published on the Agency for Healthcare Research and Quality (AHRQ) website (https://effectivehealthcare.ahrq.gov/products/interventional-treatments-pain/protocol) and on the PROSPERO systematic reviews registry (CRD42021226947). The AHRQ review examined interventional procedures for acute and chronic pain that are either not currently covered by the United States (US) Centers for Medicare & Medicaid Services (CMS), or procedures that are covered but for which there is uncertainty or controversy regarding their use, including: vertebral augmentation; cooled or pulsed radiofrequency ablation (RFA); intradiscal and facet-joint platelet-rich plasma injections; other intradiscal procedures; sphenopalatine blocks; occipital nerve stimulation; piriformis injections; and peripheral nerve stimulation. The review included 37 trials published between 2002 and 2020. ^8^

### Data Extraction

Data elements of all trials were extracted from the published articles and cross-referenced with what was reported in the AHRQ systematic review. Extracted variables included baseline characteristics of the control and intervention groups; whether baseline concurrent analgesic use was reported in the intervention and control groups; and if baseline concurrent analgesic use was reported, how it was measured and reported. We also extracted whether concurrent analgesic use was reported post-randomization in control and intervention arms, and if it was reported, how it was measured and reported. Finally, we extracted the primary pain intensity outcome at the primary trial endpoint.^10^ If cross-over was allowed, the post-randomization time point prior to cross-over was analyzed to preserve the benefits of randomization. All major data elements were extracted by two or more reviewers (BS, AB, CS, CD, MD). Areas of uncertainty were reviewed by the senior author (PS) and resolved by discussion.

### Reported concurrent analgesic use and methods used to account for concurrent analgesic use

*Baseline concurrent analgesic use variables*: If concurrent analgesic use was reported, the manner of analgesic reporting was classified as reporting of average dose of medication, prevalence of opioid use, prevalence of non-opioid use, prevalence of individual medication or medication category use, prevalence of any analgesic use, or “other”.

*Post-randomization concurrent analgesic use variables at the primary trial endpoint*: If concurrent analgesic use was reported, the manner of analgesic reporting was classified using the same options as used for classifying the baseline analgesic use reporting. If the primary trial endpoint was not explicitly noted in a trial, we decided *a priori* to define the primary endpoint (for the purposes of this meta-epidemiologic study) as the time point when the largest treatment effect was expected based on clinical knowledge, factoring in the expected onset of effect and duration of effect for both the treatment and control groups. For example, for vertebroplasty and kyphoplasty, after which pain relief is thought to be rapid, we reasoned that the maximal treatment effect would be at 1-month post-randomization. On the other hand, for RFA we reasoned that the maximal treatment effect would be at 3 months post RFA (as relief after radiofrequency is not expected to be immediate, and post-RFA periods of increased pain [“neuritis”] may occur).^11^

*Methods to account for concurrent analgesic use in the analysis of the primary pain intensity endpoint*: We classified the methods used to account for concurrent analgesic use in the analysis of the primary pain outcome, including whether analgesic use was adjusted for as a covariate, analyzed as a binary outcome, incorporated into a composite outcome, used for imputation of pain intensity, or “other“. We planned *a priori* to expand this list of categories as needed, depending on the frequency of the methods to account for concurrent analgesic use encountered among included RCTs from the AHRQ systematic review.

### Data Analysis and Synthesis

To address the descriptive primary aims of the study, we: (1) calculated the prevalence of any concurrent analgesic use reported at baseline and post-randomization, (2) calculated the frequency of different methods of reporting analgesic use, and (3) determined how frequently authors accounted for concurrent analgesic use in the analysis of pain intensity outcomes. Although the primary aims were descriptive in nature, we anticipated based on the literature ^7^ that most of the trials would not report concurrent analgesic use, that a high proportion of trials would ignore analgesic use post-randomization when analyzing pain intensity outcomes, and that the most common analytic method related to concurrent analgesic use in pain RCTs would be to analyze analgesic use as a separate binary outcome. Of note, this last method does not account for concurrent analgesic use in the analysis of pain intensity outcomes *per se* but analyzes analgesic use as a secondary outcome, ignoring that analgesic use may be impacting the primary as-randomized comparison of treatment groups with regards to pain intensity.

We also conducted a meta-analysis of treatment effects on pain intensity measured using the conventional 0 to 10 Numeric Rating Scale of pain intensity (NRS) vs. treatment effects measured using the recently-developed QPAC_1.5_ composite outcome. This meta-analysis was conducted among the subset of trials that reported conventional NRS outcomes and concurrent analgesic use (both of which are needed to calculate the QPAC_1.5_) for each of the intervention and control groups, at the primary trial endpoint. As the use of analgesics may be associated with pain intensity, when calculating the standard deviation of the QPAC_1.5_ we assumed a correlation between analgesic use and pain NRS of 0.08 based on a prior report;^6^ sensitivity analyses were conducted using other assumptions for this correlation. The details of how the QPAC_1.5_ was calculated is provided in the Supplementary Materials.

Based on the theory underlying the QPAC_1.5_, ^5^ this measure would be expected to increase the estimated treatment effect in a given RCT *if* a direct treatment effect exists when comparing the intervention and control groups and when analgesic use is differential across groups. In contrast, if no direct treatment effect exists when comparing the intervention and control treatments and when there is no treatment impact on analgesic use, then the QPAC_1.5_ would not be expected to change estimated treatment comparisons.^5^ With this reasoning in mind, we pre-specified a subset of RCTs from the AHRQ systematic review for further study in a meta-analysis comparing different methods of accounting for post-randomization concurrent analgesic use on estimated treatment effects on pain intensity. In this subset of RCTs, we expected clinically-relevant treatment effects *a priori* given the intervention and control treatments involved. Accordingly, we hypothesized that, in this subset of RCTs, analyses of the QPAC_1.5_ would produce larger treatment effects than analyses of the conventional NRS. As we have described elsewhere,^5^ the QPAC_1.5_ estimates a *controlled direct* treatment effect, whereas the conventional NRS estimates a *total* treatment effect, so each of these two measures targets a slightly different estimand. The subset of RCTs from the AHRQ review included those comparing (1) vertebral augmentation vs. usual care control, and (2) cooled RFA vs. control.^9^ We also pre-specified (3) a group of “positive control” interventional trials from outside the AHRQ review to include in the meta-analysis, in order to encompass another interventional pain treatment for which clinically-relevant treatment effects would generally be expected. For this positive control group, we selected trials of transforaminal epidural steroid injections (TF ESI) vs. control for lumbosacral radicular pain, as such trials have generally shown consistent statistically significant treatment effects on pain intensity in past RCTs.^12^ Pain intensity data for RCTs of TF ESI were extracted from a Cochrane systematic review, ^12^ and analgesic data were extracted from the original articles.

Any RCT in these 3 groups of trials which reported a pain intensity outcome at the time of the primary endpoint, and also reported concurrent analgesic use, contributed to the meta-analyses. We conducted random-effects meta-analyses. The first meta-analysis estimated the total treatment effect on pain intensity using the conventional pain intensity NRS. The second meta-analysis estimated the treatment effect on the post-randomization proportion of concurrent analgesic use. These two meta-analyses examine standard outcomes that are typically reported in pain RCTs. The third meta-analysis estimated the approximate controlled direct treatment effect on pain intensity using the QPAC_1.5._.^5^ The QPAC_1.5_ assumes that the average impact of analgesic use is a 1.5-NRS-point change in pain intensity, but otherwise estimates a well-defined contrast in mean values of a weighted composite outcome so is more generally a valid comparison.^5^ Last, the fourth meta-analysis directly assessed the impact of the QPAC_1.5_ by comparing the difference between analyses of treatment effects on pain intensity using paired analyses of the conventional pain NRS as compared to the QPAC_1.5_. Accordingly, the primary statistical inference for the secondary aim of this study was based on the fourth meta-analysis; we note that the p-value produced by this meta-analysis is equivalent to that produced by the second meta-analysis (which examined the proportion of concurrent analgesic use across the included RCTs). We considered a p-value <0.05 as the threshold for statistical significance. We examined the differences in heterogeneity (I^2^) of the meta-analyses with the conventional NRS vs. the QPAC_1.5_. Details of the meta-analyses can be found in the Supplemental Materials.

## RESULTS

The characteristics of the 37 trials included in the current review are listed in Table 1. We found that 17 of 37 (46%) of trials reported concurrent analgesic use at baseline and that the most common method used was to report the prevalence of any opioid use (11 of 37 [30%] of all trials) (Table 2).

**Table 1.**
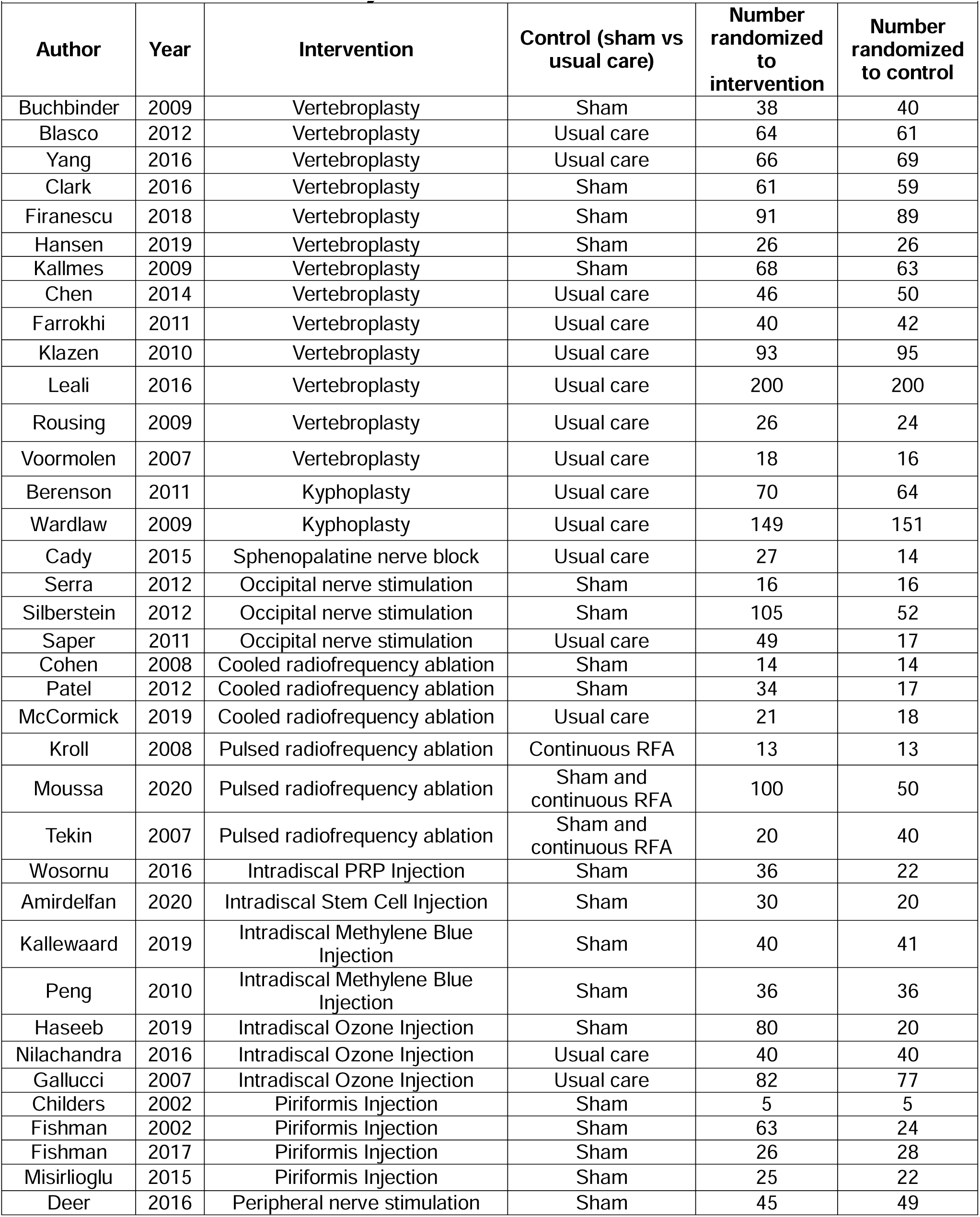
Characteristics of Analyzed Trials.

**Table 2.** Reporting of Concurrent Analgesic Use at Baseline Among Included RCTs (n=37)

|  |  |
| --- | --- |
| **Analgesic use Characteristics** | **n (%)** |
| **Reported analgesic use at baseline** (any method) | 17 (46%) |
| **Method used to report concurrent analgesia\*** |  |
| Average dose of medication | 4 (11%) |
| Prevalence of any opioid use | 11 (30%) |
| Prevalence of non-opioid use | 7 (19%) |
| Prevalence of individual medication use | 1 (3%) |
| Prevalence of any analgesic use | 7 (19%) |
| Other method | 1 (3%) |

\*Some studies reported more than one method

We also found that 20 of 37 (54%) of the studied trials reported concurrent analgesic use post-randomization and that the most common method used was to report the prevalence of any analgesic use (11 of 37 (30%) of all trials) (Table 3). Four of 37 (11%) of trials used analgesic scoring methods that were specific to their trial and not otherwise widely used in the field. For example, one trial created an arbitrary grading for each type and/or dose of analgesic and summed the gradings across analgesics used.^13^ The most common method of incorporating analgesic use in analyses was to report analgesic use as a binary outcome. Fourteen of 37 (38%) of trials analyzed whether the pain intervention affected the frequency of analgesic use. Only one trial accounted for concurrent analgesic use directly in their analysis of treatment effects on pain intensity, by using a binary composite “responder” outcome. This responder outcome was defined as a 30% pre-to-post reduction in pain intensity without an increase in pain medication.^14^ None of the included trials accounted for analgesic use using the approach of treating the pain NRS as if it was missing when there is concurrent analgesic use, a strategy that is sometimes used.^10^

**Table 3.** Reporting on Concurrent Analgesic Use Post-Randomization Among Included RCTs (n=37)

|  |  |
| --- | --- |
| **Analgesic use Characteristics** | **n (%)** |
| **Reported analgesic use post-randomization** (any method) | 20 (54%) |
| **Method used to report concurrent analgesia** |  |
| Average dose of medication | 1 (3%) |
| Prevalence of any opioid use | 4 (11%) |
| Prevalence of non-opioid use | 3 (8%) |
| Prevalence of individual medication use | 1 (3%) |
| Prevalence of any analgesic use | 11 (30%) |
| Other method | 4 (11%) |

Only 5 trials from the AHRQ systematic review met our pre-specified inclusion criteria (intervention-control comparisons for which clinically relevant treatment effects were expected *a priori*) and had the requisite data needed to calculate the QPAC_1.5_ in treatment and control groups (pain intensity outcome reporting and concurrent analgesic use prevalence at the primary pain intensity endpoint), and therefore were included in the meta-analyses.^15–19^ Four meta-analyses were conducted in this subgroup of 5 trials. Among the trials of transforaminal lumbar epidural steroid injections from the Cochrane review that we considered for inclusion in the meta-analysis (the “positive control” group),^12^ none reported concurrent analgesic use at the primary endpoint when pain intensity outcomes were reported. Therefore, the QPAC_1.5_ could not be calculated for these studies and they were not included in the meta-analyses.

In the first meta-analysis, we examined treatment effects on the conventional pain intensity NRS at the primary trial endpoint in the 5 trials that met our inclusion criteria. This revealed a significant (beneficial) average treatment effect on pain intensity comparing the interventional treatments to controls of –2.31 NRS points (95% CI –3.50 to –1.11, Supplemental Figure 1), reflecting less pain in those who received interventional pain treatments. In the second meta-analysis, we estimated the average treatment effect on the post-randomization proportion of concurrent analgesic use. We found a statistically significant 30% lower proportion of analgesic use in patients who received the interventional treatments (p<0.001), as compared to the control treatments (Supplemental Figure 2).

In the third meta-analysis, we estimated treatment effects on pain intensity measured using the QPAC_1.5_. We found a large-magnitude treatment effect for interventional pain treatments as compared to control treatments (Supplemental Figure 3), with an estimated difference of –2.77 points on the QPAC_1.5_ (95% CI –4.17 to –1.37). This treatment effect point estimate (−2.77 points) was slightly larger than the treatment effect on the pain intensity NRS (−2.31 points) found in the first meta-analysis. One trial of the 5 included in the meta-analysis, which had not show a statistically significant improvement with the interventional treatment when analyzing the conventional pain intensity NRS (Supplemental Figure 1), *did* show a statistically significant improvement with the interventional treatment when analyzing the QPAC_1.5_.^16^ Finally, in the fourth meta-analysis, on which we based our primary statistical inference, we estimated the difference between treatment effects using the conventional pain NRS as reported and treatment effects using the QPAC_1.5_. We found a significantly larger overall treatment effect with the QPAC_1.5_ as compared to the standard analysis of the conventional NRS while ignoring concurrent analgesic use, with a reduction in pain intensity of −0.45 NRS points [95% CI −0.76 to −0.14]; p<0.001 (Figure 1). This indicates a nearly 0.5 NRS-point larger treatment effect on pain intensity when accounting for concurrent analgesic use via the QPAC_1.5_. Heterogeneity with all four analyses was high (85-90%).

**Figure 1.**
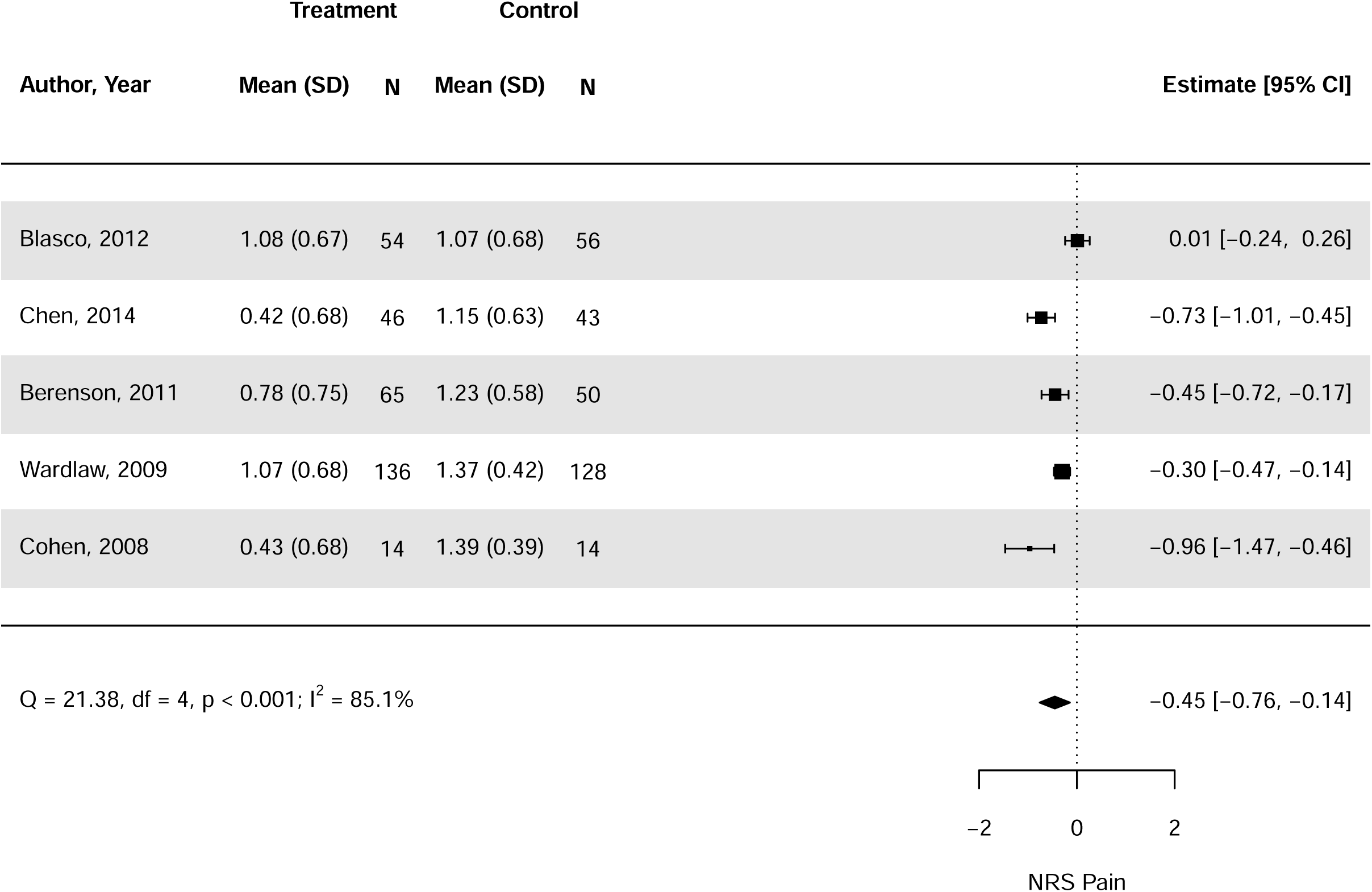
Forest plot of the difference in the treatment effect on the conventional pain intensity NRS (ignoring concurrent analgesic use) vs. the treatment effect on the QPAC_1.5_ (n=5 trials) Point estimates less than 0 indicate that the treatment effect on pain intensity using the QPAC_1.5_ is, on average, larger than the treatment effect on pain intensity using the conventional pain NRS. Whiskers show the bounds of the 95% confidence intervals.

Sensitivity analyses assuming different correlations between concurrent analgesic use and pain NRS scores, and using leave-one-out meta-analyses, did not show material differences from the primary results in terms of treatment effect estimates or heterogeneity (data not shown).

## DISCUSSION

This study sought to determine the frequency of reporting concurrent analgesics use at baseline and post-randomization among interventional pain RCTs included in a high-quality systematic review of interventional procedural treatments for pain. We also conducted a meta-analysis to determine if accounting for concurrent analgesic use in analyses of pain intensity using a novel composite pain intensity-analgesia outcome, the QPAC_1.5_, would produce a significantly larger treatment effect size than analyses of the conventional pain NRS. We found that only half of the trials studied reported whether concurrent analgesics were used at baseline and post-randomization, and only one trial directly accounted for concurrent analgesic use post-randomization in the primary pain intensity outcome.^14^ Our meta-analysis found that accounting for concurrent analgesic use with the QPAC_1.5_ led to statistically significantly larger treatment effects on pain intensity of –0.45 NRS points (95%CI –0.76 to –0.14) compared to analyses of pain intensity using the conventional NRS.

Randomized controlled pain trials are very important for determining the effectiveness of new interventional pain medicine treatments. Recent work suggests that a seemingly minor secondary outcome reported in pain RCTs, concurrent analgesic use, can have a substantial impact on overall treatment effect estimates, sometimes affecting whether a trial yields a statistically significant result for the primary pain intensity endpoint.^6^ The current study highlights that only half of interventional pain trials studied in a major systematic review reported concurrent analgesic use. Moreover, when pain trials do assess and report concurrent analgesic use, they usually report it as the prevalence of any analgesic used and do not directly account for this use in their primary outcomes. Assessing analgesic use as a binary outcome does not allow investigators to account for analgesic use in the analyses of treatment effects on pain intensity, but treats analgesic use as a distinct outcome, when in actuality both pain intensity and analgesic use outcomes reflect different aspects of the same pain construct (improvement in pain intensity). This can alter treatment effect estimates and potentially fundamentally affect the main conclusions drawn from a pain RCT. A unique finding from the current study is that accounting for concurrent analgesic use when analyzing pain intensity outcomes using the QPAC_1.5_ increased treatment effect estimates on pain intensity by nearly 0.5 NRS points, indicating a larger *beneficial* effect of the procedural treatments studied. One out of 5 trials in the meta-analysis shifted from having non-significant to significant effects on pain intensity after applying the QPAC_1.5_.^16^ This illustrates how accounting for analgesic use and pain intensity in the same composite outcome can potentially improve the power of pain RCTs and even affect the main conclusions drawn from certain trials. This finding is consistent with prior work,^5^ supporting the composite score’s validity.

The current study’s findings reinforce the need to uniformly report and account for analgesic use in interventional pain trials. The pre-specified subset of RCTs we selected for the current meta-analysis was chosen specifically because they included treatment vs. control contrasts where large treatment effects on pain intensity were expected. This is the context in which larger between-group differences in analgesic would be expected, resulting in larger estimates of the treatment effects on pain intensity when using the QPAC_1.5_. It cannot be expected that the QPAC_1.5_ will always increase treatment effect size estimated on pain intensity when applied to RCT data, however, and in the context of an RCT of a treatment with no meaningful treatment effect on pain intensity, the QPAC_1.5_ would not be expected to produce a net increase in treatment effect size.

One implication of this study is that a lack of accounting for concurrent analgesic use may increase the proportion of trials not finding statistically significant between-group differences in treatment effects and may decrease the size of the estimated treatment effects such that clinically relevant treatment effects are interpreted as not being clinically relevant.^20^ This may carry forward into evidence synthesis and clinical practice guidelines, resulting in potentially effective pain treatments being discarded. This possibility suggests that further studies of methods to account for concurrent analgesic use in pain RCTs should perhaps be a high priority for future research.

A strength of the current work is that it is the first comprehensive study to examine the frequency of reporting of concurrent analgesic use and the impact of analgesic use on the overall treatment effects of interventional pain trials. Limitations include the fact that we included clinical trials from only one systematic review of interventional pain trials from 2021, which focused primarily on emerging interventional treatments. These findings therefore may not represent other trials of other pain treatments, nor the current state of pain trials reported in the last 4 years. While responder outcomes were not examined in this study, as the QPAC_1.5_ is oriented towards analysis of mean outcomes and the same responder outcomes were not reliably reported across studies (i.e., not permitting meta-analysis), the QPAC_1.5_ approach could be converted into a binary responder outcome. Furthermore, heterogeneity was high in the meta-analyses we conducted, but was consistently high with or without applying the QPAC_1.5_. We used a random-effects meta-analysis to account for this, yet despite the often lower power of this meta-analytic approach, we still found statistically significant results. Nevertheless, future studies are needed to evaluate the applicability of the current findings to trials of other procedural and non-procedural pain treatments.

## CONCLUSIONS

This study found that about half of non-surgical interventional pain RCTs from a major systematic review did not report on post-randomization concurrent analgesic use, and that only one trial accounted for analgesic use directly in analyses of treatment effects on pain intensity at the primary trial endpoint. When accounting for concurrent analgesic use with the QPAC_1.5_ in a meta-analysis, treatment effects on pain intensity were nearly 0.5 NRS points larger compared to when concurrent analgesic use was ignored. Future studies are needed to determine how pain trials should best account for concurrent analgesic use in the analyses of pain intensity outcomes. Applying those methods to pain RCTs may improve our ability to identify treatments with clinically relevant effects.

## Data Availability

All data reported in the present work are already publicly available in previously published research articles or contained in the manuscript.

## ACKNOWLEDGEMENTS

We thank Coralynn Sack, MD, MPH for providing feedback on the final manuscript.

