## Supplemental Materials for "Methods used to Account for Concurrent Analgesic Use in Randomized Controlled Trials of Interventional Pain Treatments: A Meta-Epidemiologic Study"

This document contains one section of “Supplemental Methods” and one section of “Supplemental Results”.

**Supplemental Methods**

**QPAC_1.5_ calculation**

The quantitative pain and analgesic composite outcome or QPAC_1.5_ is a composite outcome measure that was developed to impute a counterfactual pain intensity outcome for participants who have recently used analgesics: what their pain intensity *would have been* had they not taken analgesics. Further detail regarding the use of the measure in analyses can be found elsewhere.[5]

In groups of people, the counterfactual pain intensity outcome can be calculated as QPAC_1.5=_ $\bar{X}+1.5p_{a}$ , where $\bar{X}$ is the primary pain intensity outcome at the primary pain intensity endpoint and $p_{a}$ is the proportion of concurrent analgesic use at the primary pain intensity endpoint. This resulting value is then the new pain intensity mean value that can be used in the meta-analysis. Further information is provided below.

**Meta-analysis calculation**

In order to conduct the meta-analyses, the mean and standard deviation of the outcome measure are needed for both the treatment and control arms. Below, we demonstrate how this information was obtained for each meta-analysis.

**Meta-analysis of pain NRS scores as reported.**

All values used in this analysis were taken directly from the 5 published studies included in the 2021 AHRQ systematic review [8] that met pre-specified inclusion criteria (intervention-control comparisons for which clinically relevant treatment effects were expected *a priori*). [16-20] For subsequent calculations, we use the following notation. We do not distinguish between the treatment and control groups here however each value was evaluated separately for each group.

- Mean pain NRS score reported in literature: $\bar{X}$
- Standard deviation of the pain NRS score reported in literature: $\hat{SD}$
- Proportion of concurrent analgesic users reported in the literature: $p_{a}$

**Meta-analysis of pain NRS scores using QPAC_1.5_**

The values in this analysis were derived using the mean and standard deviation of pain NRS scores as well as the proportion of concurrent analgesics users in the literature. Additionally, an estimate of the correlation between concurrent analgesic use and pain NRS scores was obtained from the literature (0.08). Because the true correlation is unknown, two other correlations were assumed in sensitivity analyses: a correlation of 0 , and a two-fold larger correlation of 0.16. The standard deviation was calculated using the equation for the variance of the sum of two dependent variables.

- Mean pain NRS score using QPAC_1.5_: $\bar{X}+1.5p_{a}$
- Standard deviation of the pain NRS score reported in literature: $\sqrt{\hat{SD}^{2}+2.25p_{a}\left( 1-p_{a} \right)+3\rho\hat{SD}\sqrt{p_{a}(1-p_{a})}}$

Where $\rho$ denotes the correlation between concurrent analgesic use and pain NRS scores.

**Meta-analysis comparing pain NRS scores from published studies [16-20] and pain NRS scores using QPAC_1.5_**

For these equations we calculated the difference between the reported and QPAC_1.5_ pain NRS scores in each treatment group which simplifies to the proportion of concurrent analgesic users multiplied by 1.5

- Mean difference between QPAC_1.5_ and reported NRS scores: $1.5p_{a}$
- Standard deviation of the difference between QPAC_1.5_ and reported NRS scores: $1.5\sqrt{p_{a}(1-p_{a})}$

Sensitivity analyses for the above meta-analyses were conducted using leave-one-out analyses.

**Supplemental Results**

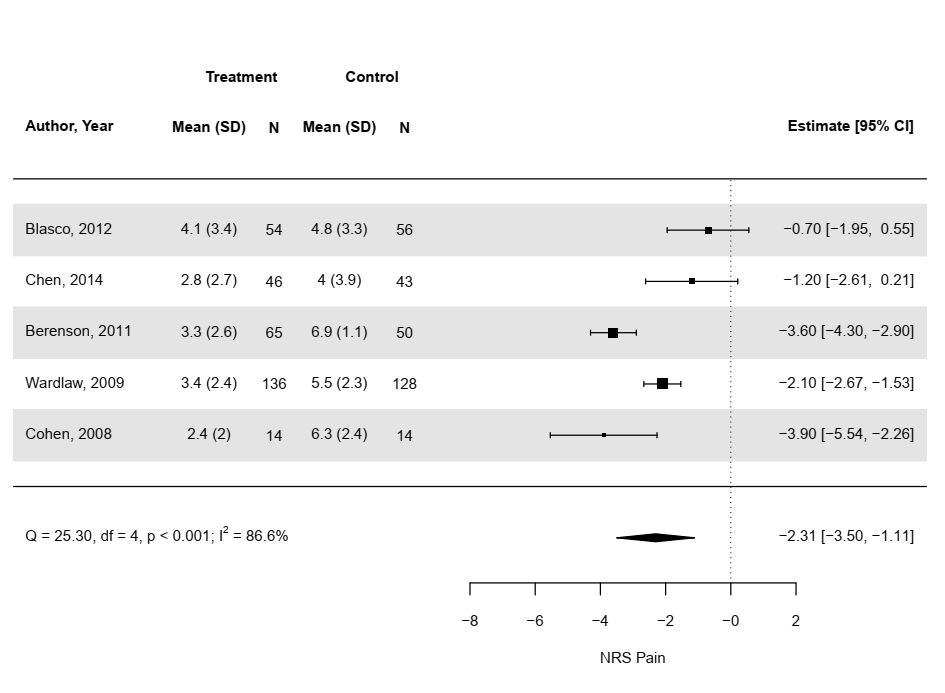

Supplemental Figure 1. Forest plot of treatment effects from included RCTs, analyzing the conventional pain intensity NRS at the primary pain intensity endpoint as the outcome, ignoring concurrent analgesic use.

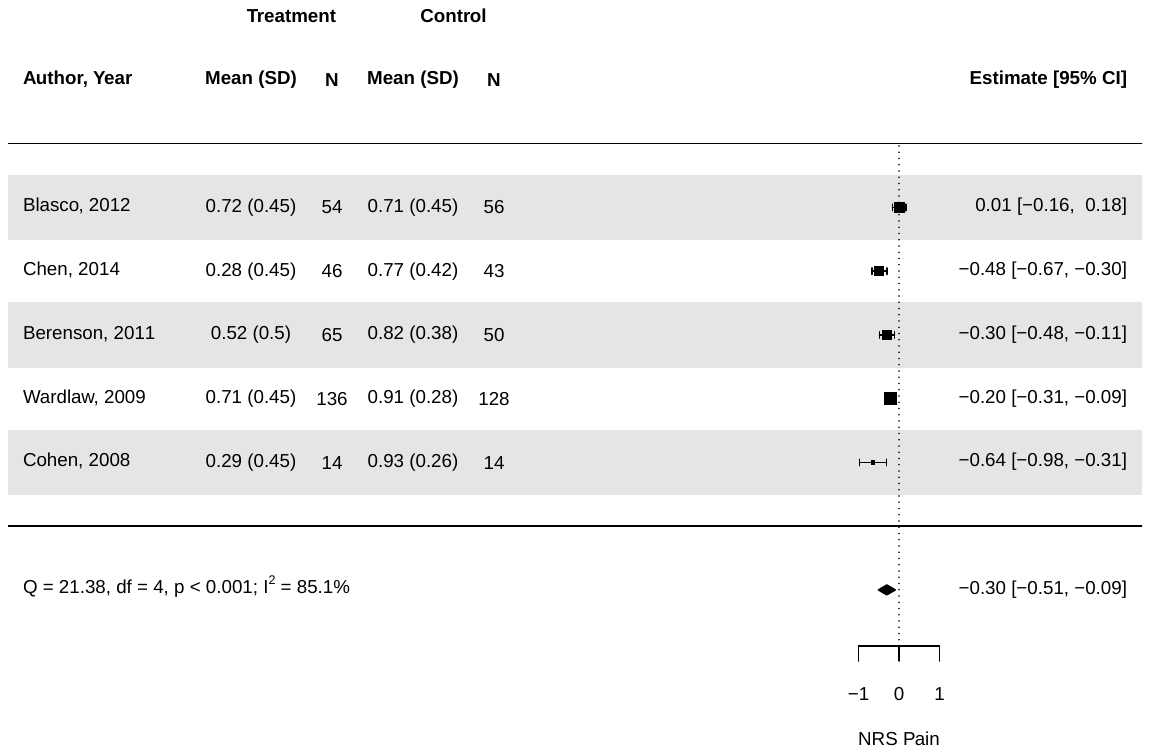

Supplemental Figure 2. Forest plot of treatment effects from included RCTs, analyzing post-randomization analgesic use at the primary trial endpoint as the outcome.

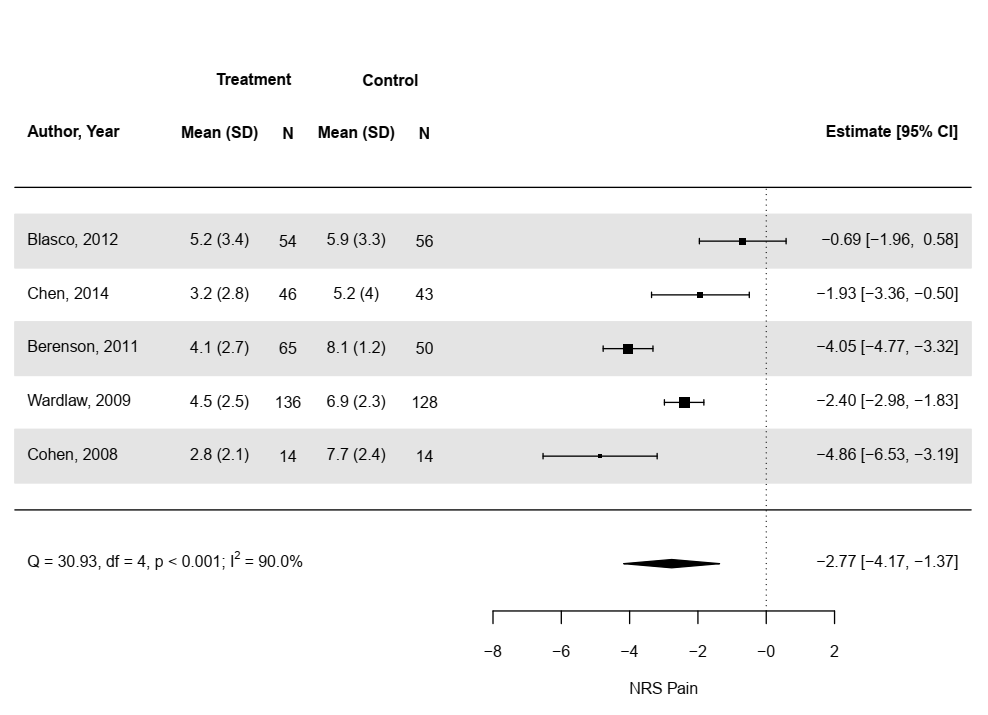

Supplemental Figure 3. Forest plot of treatment effects from included RCTs, analyzing the QPAC_1.5_ at the primary pain intensity endpoint as the outcome.
